# Elevated HbA1c is associated with advanced brain age in severe obesity

**DOI:** 10.64898/2026.06.04.26354935

**Authors:** Joshua Juhasz, Brittany DeFeis, Mark K. Britton, Hannah Hoogerwoerd, Kate Worwag, Keyanni J. Johnson, Angel Uribe, John B. Williamson, Eric C. Porges, Ronald A. Cohen

## Abstract

**Introduction:** Brain-predicted age, estimated from structural MRI data, is a machine-learning biomarker of biological brain aging. Greater brain age gap (BAG) indicates advanced brain aging and is associated with cognitive decline and mortality. Cardiometabolic risk factors, including elevated blood glucose, body mass index (BMI), blood pressure, and cholesterol, increase risk of cognitive impairment and dementia in aging. Their relationship with BAG in severe obesity remains poorly characterized despite increased prevalence of cardiometabolic risk factors among this population.

**Methods:** T1-weighted MRI data from 97 adults (BMI 35-73) were used to calculate BAG using ENIGMA and Pyment brain age models. Associations between BAG and HbA1c, BMI, hypertension, and hyperlipidemia were examined using multiple linear regression and MM–estimation robust regression, adjusting for age, sex, and race. Post hoc analyses stratified models by clinical HbA1c cutoffs (normoglycemic, prediabetic, diabetic).

**Results:** Higher HbA1c was associated with greater BAG_ENIGMA_ (*B* = 1.58, *p* = .014) and BAG_Pyment_ (*B* = 0.93, *p* = .013) in linear regression models. In robust models, HbA1c remained significantly associated with BAG_ENIGMA_ (*B* = 1.70, *p* = .002) but not BAG_Pyment_ (*B* = 0.71, *p* = .13). BMI, hypertension, and hyperlipidemia were not associated with BAG in either linear or robust models. HbA1c was associated with greater BAG_ENIGMA_ (*B* = 2.15, *p* = .01) and BAG_Pyment_ (*B* =1.21, *p* = .04) in those at or above prediabetic levels and with BAG_ENIGMA_ (*B* = 2.49, *p* = .047) in those with diabetes.

**Conclusions:** Elevated HbA1c is associated with advanced brain aging in individuals with severe obesity. BAG was not associated with BMI, hypertension, and hyperlipidemia, which may reflect the restricted BMI range inherent to the sample with severe obesity.

**HIGHLIGHTS:**

- Elevated HbA1c was associated with advanced brain aging in individuals with severe obesity.
- HbA1c-BAG associations were seen primarily in the context of clinically elevated levels (i.e. prediabetes and diabetes).
- The study did not provide evidence to support that BMI, hypertension, and hyperlipidemia are associated with BAG in individuals with severe obesity
- ENIGMA and Pyment models showed limited agreement in BAG estimates in a sample of individuals with severe obesity.

## 1. INTRODUCTION

### 1.1 Brain-Predicted Age and Brain Age Gap

The brain undergoes structural changes during aging, including cortical thinning, brain volume loss, and increased white matter hyperintensities.^1,2^ Each of these changes have been found to be associated with decreases in cognition and neurological function.^3–10^ Brain-predicted age, or brain age, is a biomarker of structural and biological aging processes.^11–13^ It uses machine-learning algorithms trained on large datasets of healthy individuals to estimate age, derived from T1-weighted MRI images or brain volume, surface area, and thickness measurements.^11–14^ Brain age gap (BAG), also called brain predicted age difference, reflects an individual’s brain-predicted age minus the actual age of an individual.^11–16^ More positive values (where actual age is less than predicted) reflect biologically older, less healthy brains.^11^ More negative values (where actual age is greater than predicted) reflect biologically healthier, younger brains.^11^ Increased BAG has been associated with increased hazard of mild cognitive impairment, dementia, and mortality.^11,12^ Several publicly available brain age algorithms have been used to generate brain-predicted age as a biomarker of biological brain aging.^14^ BAG has also been used to find associations between advanced and accelerated biological aging of the brain and neurological disorders, metabolic disorders, cardiovascular conditions, psychiatric disorders, and lifestyle factors.^11–17^

### 1.2 Cardiometabolic Risk Factors and Metabolic Syndrome

Metabolic syndrome (Met-S), a cluster of cardiovascular and metabolic conditions is associated with a 2-fold increase in relative risk of cardiovascular disease (e.g., stroke and myocardial infarction).^18,19^ Met-S is characterized by dyslipidemia, elevated triglycerides, elevated blood glucose hypertension, and abdominal obesity.^18^ Met-S, as well as individual tests for elevated blood glucose, hypertension, hyperlipidemia, and obesity (body mass index ≥ 30) have been shown to be associated with worsened brain health outcomes as individuals age, such as mild cognitive impairment, dementia, and progression of cognitive impairment to dementia.^19–25^ Cardiometabolic risk can also be characterized by factors and measurements not included in the diagnostic criteria of metabolic syndrome (e.g., HbA1c, BMI, elevated total cholesterol), which have also been shown to correlate with increased risk of cardiovascular disease outcomes and worsened brain health in aging.^23,25–30^ Cardiometabolic risk factors are more prevalent in individuals with obesity, demonstrating increased risk and importance in understanding cardiometabolic risk among individuals with obesity.^18,19^ Notably, many of these risk factors are modifiable with lifestyle change, pharmacological or surgical interventions.^31^ Their relationship to health outcomes can been used as justification for modifying cardiometabolic risk, when possible, to invoke health benefits.^31^

Obesity can be classified into class I, II, or III using body mass index (BMI) values between 30 and 35, 35 and 40, and ≥40, respectively.^32^ Severe obesity is defined as having class III obesity or class II obesity with at least one obesity-related comorbidity.^33^ BAG is reported to be associated with cardiometabolic risk factors in healthy populations and populations with obesity.^27–30^ However, literature investigating the associations between cardiometabolic risk factors and BAG is sparse in populations with severe obesity.^30^

More insight into the relationship between these modifiable cardiometabolic risk factors and the difference between brain-predicted age and chronological age for those with severe obesity would provide insights into whether additional cardiometabolic risk factors are related to advanced aging in populations who already have obesity. This could elucidate the need for interventions not only targeting weight loss but also focusing on specific lifestyle interventions to reduce risk related to brain health as this population ages. Testing the reliability of two open-source brain age algorithms could also serve to discern the accuracy and reliability of these algorithms for clinical use, specifically in populations with severe obesity.

This study aims to 1) investigate the cross-sectional relationships between several cardiometabolic risk factors (hypertension, hyperlipidemia, BMI, and HbA1c) and brain health in aging, using brain age gap as a biomarker, and 2) compare ENIGMA and Pyment model brain-predicted age estimation, within a sample of individuals with severe obesity. We hypothesized that BMI, HbA1c, hypertension, and hyperlipidemia would each be associated with increased BAG from both models, representing less healthy brain aging. We also hypothesized that the ENIGMA and Pyment models would generate similar, and not systematically biased BAG values.

## 2. METHODS

### 2.1 Sample

The sample in this study includes 97 individuals with severe obesity; individuals had either class II (BMI ≥ 35) obesity and a significant medical condition related to obesity (e.g., type II diabetes, sleep apnea, or heart disease) or class III (BMI ≥ 40) obesity. Sample demographics, including age, years of education, race, and sex, are included in **Table 1**. Participants were recruited as part of a larger clinical trial examining changes in cognition following bariatric surgery (R01DK099334) and included both individuals who planned on undergoing bariatric surgery at the University of Florida or as community dwelling matched controls. All participants met University of Florida clinical requirements for bariatric surgery (e.g., Roux-en-Y procedure) and provided written informed consent to participate in the study. Study procedures were approved by the University of Florida IRB (IRB201400034) and conformed to the Declaration of Helsinki.

**Table 1.** Descriptive Statistics for Cardiometabolic, Demographic, and Brain Age Variables.

| Characteristic | Mean | Standard Deviation | % of Characteristic |
|---|---|---|---|
| Age | 44.7 | 12.3 | -- |
| Race (White) | -- | -- | 67.0 |
| Sex (Female) | -- | -- | 80.4 |
| BMI | 46.3 | 8.0 | -- |
| Years of Education | 14.0 | 2.3 | -- |
| HbA1c | 6.2 | 1.3 | -- |
| Hypertension | -- | -- | 59.8 |
| Hyperlipidemia | -- | -- | 34.0 |
| ENIGMA Brain Age | 48.4 | 11.7 | -- |
| Pyment Brain Age | 42.3 | 13.0 | -- |
| BAG<sub>ENIGMA</sub> | 3.7 | 9.0 | -- |
| BAG<sub>Pyment</sub> | -2.3 | 4.4 | -- |

Descriptive statistics for cardiometabolic variables, medical variables, brain predicted-age and brain age gap are shown.

### 2.2 Cardiometabolic Risk Factor Measurements and Collection

HbA1c, or glycated hemoglobin, is a biomarker for elevated blood glucose, measuring the average blood glucose of an individual over the last three months.^34^ Values below 5.7% are considered normal, values between 5.7% and 6.4% indicate prediabetes, and values of 6.5% or higher meet the diagnostic criteria for diabetes mellitus.^34^

Body Mass Index (BMI), weight (kg) divided by height squared (m^2^) was used to categorize participants as having class II obesity (35.0 to 39.9 kg/m^2^) and class III obesity (≥40.0 kg/m^2^) according to the current WHO guidelines.^31^

HbA1c was collected from blood tests at baseline participant visits as a measure for elevated blood glucose and was collected for 95.9% of the participants. Body Mass Index (BMI) was measured for 100% of participants. Lifetime self-report hypertension and hyperlipidemia data were collected for 100% of participants as answers “Yes” or “No” to ever having either condition.

### 2.3 MRI Acquisition

Structural T1-weighted images were acquired on a 3T Siemens Verio scanner (Siemens Healthcare, Erlangen, Germany) at UF Health Gainesville using a 12-channel Head Matrix coil. A 3D T1-weighted magnetization-prepared rapid gradient-echo (MPRAGE) sequence was used with the following parameters: TR = 2530 ms, TE = 3.5 ms, TI = 1100 ms, flip angle = 7°, and 1.0 mm slice thickness. The acquisition matrix was 256 × 256 with a 100% phase FOV, resulting in a voxel size of 1.0 × 1.0 × 1.0 mm. Parallel imaging was employed using a GRAPPA factor of 2. Data were converted from DICOM to NIfTI format using dcm2niix (v1.0.20180622).

### 2.4 Brain Age Algorithms

After MRI data collection, T1-weighted images were processed using FreeSurfer 6.0.0 to extract brain volume, cortical thickness, and surface area metrics using the Desikan-Killiany atlas.^35,36^ All images were visually inspected to assess quality prior to analysis. Likewise, FreeSurfer output was visually inspected for quality control; manual editing was performed, when necessary, by members of the research team. The derived structural measurements were subsequently input into the open-source ENIGMA brain age models (separate models for male and female participants), while the raw T1-weighted MRI images were analyzed using the Pyment brain age model. Both models generated brain age estimates. Both models were included due to differences in training datasets, data processing level, and mean absolute error for BAG on testing datasets. Including both models helps to better understand the reliability of brain-predicted age estimates in clinical populations across different algorithms. BAG was calculated for each participant by subtracting their chronological age from the estimated brain age produced by each model.

The ENIGMA brain age metric was chosen due to previous use in the literature in clinical populations, specifically with psychiatric conditions including major depressive disorder.^17^ It includes two algorithms, one for each sex. The models use ridge regression from the python-based “scikit-learn” package to estimate age from extracted structural measurements from FreeSurfer software.^17^ The algorithms use 77 brain structural features averaged across right and left hemispheres (including subcortical volume, cortical thickness, and cortical surface area). These measurements used in training the model were derived from FreeSurfer 5.1, 5.3, and 6.0.^17^ The model for males was trained on 952 healthy controls and the model for females was trained on 1236 healthy controls from 21 international datasets.^17^ When testing for associations with chronological age, Han et al. reported that the brain age estimates had a mean absolute error of 6.50 years for males and 6.85 years for females and Pearson’s *r* of .85 for both males and females on a testing dataset healthy controls.^17^

The Pyment brain age model uses a simple fully convolutional neural network (SFCN) to derive brain age from T1-weighted MRI images.^37^ The Pyment implementation used here implements a regression variant created by Leonardsen et al.,^37^ which was based on a previously created model with a SoftMax output.^38^ The model has one output node that predicts a continuous value representing age.^37^ The model optimizes Mean Squared Error (MSE) during training, which penalizes large deviations between predicted and actual ages.^37^ The model was trained on 34,285 individuals aged 3-95 from 21 international datasets.^37^ When testing for associations with chronological age, Leonardsen et al.^37^ reported that the brain age estimates a mean absolute error of 3.9 years and Pearson’s r of .975 on a testing dataset of healthy controls.^37^

### 2.5 Statistical Analyses

Analyses were performed in R version 4.5.0.^39^. Descriptive statistics including mean age, BAG, BMI, HbA1c, and years of education were collected for the sample. Due to non-normal distributions, Spearman correlations and Wilcoxon rank-sum tests were used to assess associations between BMI and participant characteristics (HbA1c, self-report hypertension, self-report hyperlipidemia, race, years of education, sex, and age). Two multiple linear regression models (separate for Pyment and ENIGMA algorithms) tested for associations between BAG and BMI, HbA1c, self-report hypertension, self-report hyperlipidemia, race, years of education, sex, and age.

Cook’s Distance for each linear regression model was calculated using 4/n, where n is the number of individuals in the sample, as an indicator of outliers.^40^ Residual vs. Leverage Plots were also created using 2p/n, where p is the number of predictors and n is the number of individuals in the sample, to indicate high leverage points.^40^ Cook’s distance and residual vs leverage plots are included as **Supplementary Figure 1**. Robust linear regression models using MM-estimation to down-weigh points with high leverage were fit using robustbase 0.99-66.^41^ These models tested for associations between BAG and BMI, HbA1c, self-report hypertension, self-report hyperlipidemia, race, years of education, sex, and age. A Bland-Altman test was conducted using the blandr R package 0.6.0 to test the agreement between BAG for the ENIGMA and Pyment models.^42^

Post-hoc MM-estimation robust models stratifying by HbA1c-defined glycemic status (normal/prediabetic/diabetic) were run. The models estimated BAG including HbA1c, BMI, hypertension, hyperlipidemia, race, age, and sex in the models.

## 3. RESULTS

### 3.1 Descriptive Statistics

The study sample was mostly female and White. On average, they were middle-aged and received some post-secondary education. Participants had prediabetic HbA1c levels on average, and the majority of participants had hypertension. Notably, average brain age estimates for ENIGMA were higher than chronological age, while Pyment estimates were lower than chronological age. Participant demographics, clinical characteristics, brain-predicted age and brain age gap values are summarized in **Table 1**.

### 3.2 Cardiometabolic Risk Factor Associations with Demographic Variables

Bivariate associations between cardiometabolic risk factors and demographic variables were evaluated as potential demographic confounders of the relationships between cardiometabolic risk factors and BAG. BMI, HbA1c, hypertension, and hyperlipidemia were significantly associated with age. Hypertension was also associated with race, with a higher proportion of White participants in the hypertensive group compared to the nonhypertensive group. No other significant bivariate associations between cardiometabolic risk factors and demographic variables were found. **Tables 2 and 3** and **Figures 1** show bivariate associations and test characteristics of demographic variables with BMI, HbA1c, hypertension and hyperlipidemia, respectively.

**Table 2.** Bivariate Associations of BMI, HbA1c, and Demographic Variables.

| *BMI* |  |  |  |  |
|---|---|---|---|---|
| Characteristic | Wilcoxon U | Wilcoxon r | Spearman’s ρ | *p* |
| Sex (Female) | 724 | .02 | -- | .88 |
| Age | -- | -- | -.29 | < .01* |
| Years of Education | -- | -- | -.12 | .25 |
| Race (White) | 1176 | .12 | -- | .24 |
| HbA1c |  |  |  |  |
| Characteristic | Wilcoxon *U* | Wilcoxon *r* | Spearman’s *ρ* | *p* |
| Sex (Female) | 645 | .06 | -- | .58 |
| Age | -- | -- | .42 | < .01* |
| Years of Education | -- | -- | -.06 | .57 |
| Race (White) | 1010 | .07 | -- | .50 |

Demographic variable associations are described using statistical characteristics. Categorical variables are represented as n (%); numerical variables represented as Mean (SD). Pearson’s Chi-squared test was used for categorical variable associations. Wilcoxon Rank Sum Test was used for numerical variable associations.

**Table 3.**
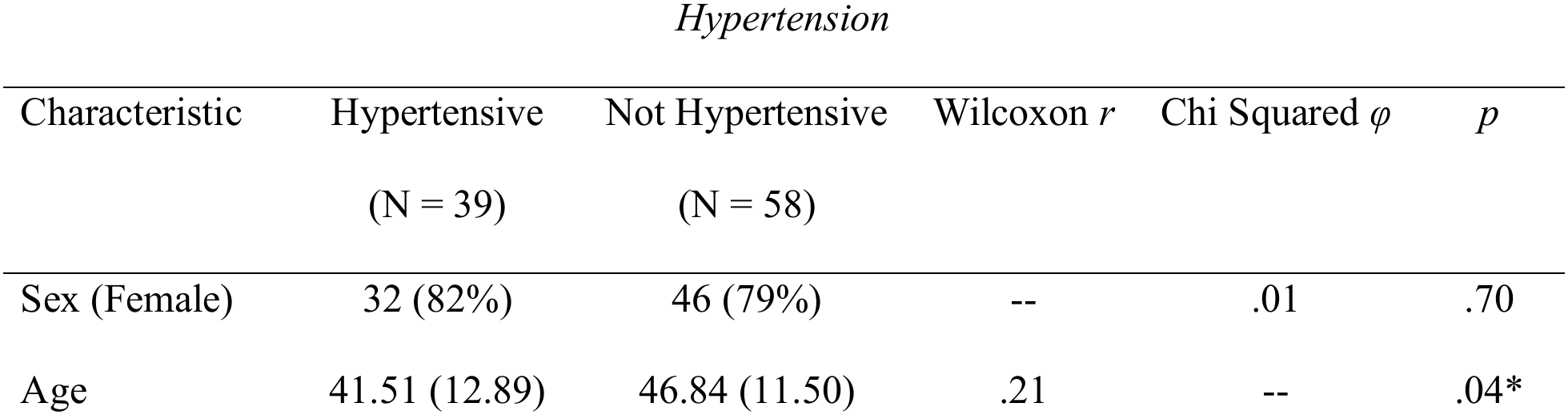
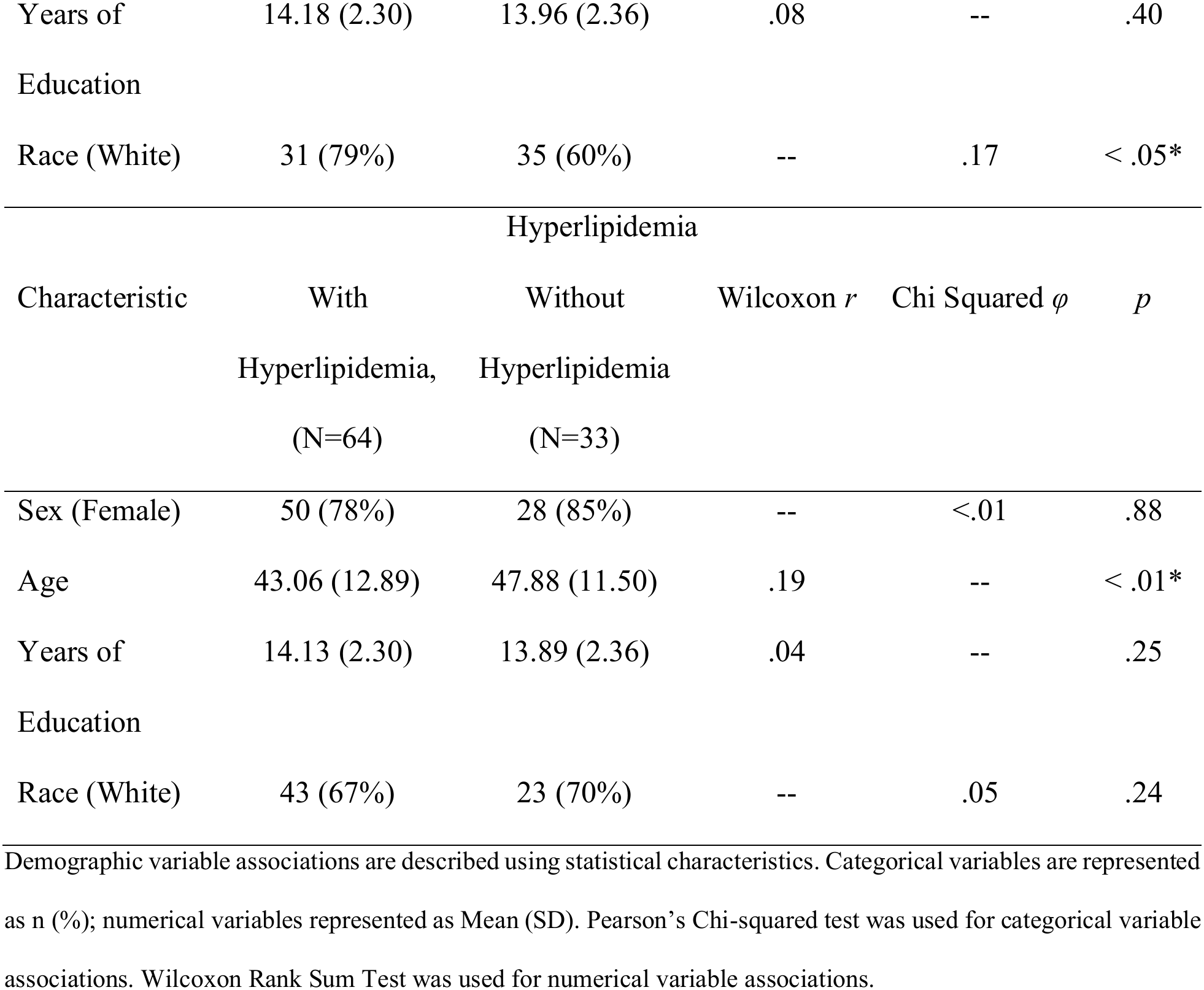
Bivariate Associations of Hypertension and Hyperlipidemia with Demographic Variables.

| *Hypertension* |  |  |  |  |  |
| Characteristic | Hypertensive (N = 39) | Not Hypertensive (N = 58) | Wilcoxon *r* | Chi Squared *φ* | *p* |
|---|---|---|---|---|---|
| Sex (Female) | 32 (82%) | 46 (79%) | -- | .01 | .70 |
| Age | 41.51 (12.89) | 46.84 (11.50) | .21 | -- | .04* |
| Years of Education | 14.18 (2.30) | 13.96 (2.36) | .08 | -- | .40 |
| Race (White) | 31 (79%) | 35 (60%) | -- | .17 | < .05* |
| Hyperlipidemia |  |  |  |  |  |
| Characteristic | With Hyperlipidemia, (N=64) | Without Hyperlipidemia (N=33) | Wilcoxon *r* | Chi Squared *φ* | *p* |
| Sex (Female) | 50 (78%) | 28 (85%) | -- | <.01 | .88 |
| Age | 43.06 (12.89) | 47.88 (11.50) | .19 | -- | < .01* |
| Years of Education | 14.13 (2.30) | 13.89 (2.36) | .04 | -- | .25 |
| Race (White) | 43 (67%) | 23 (70%) | -- | .05 | .24 |

Demographic variable associations are described using statistical characteristics. Categorical variables are represented as n (%); numerical variables represented as Mean (SD). Pearson’s Chi-squared test was used for categorical variable associations. Wilcoxon Rank Sum Test was used for numerical variable associations.

**Figure 1.**
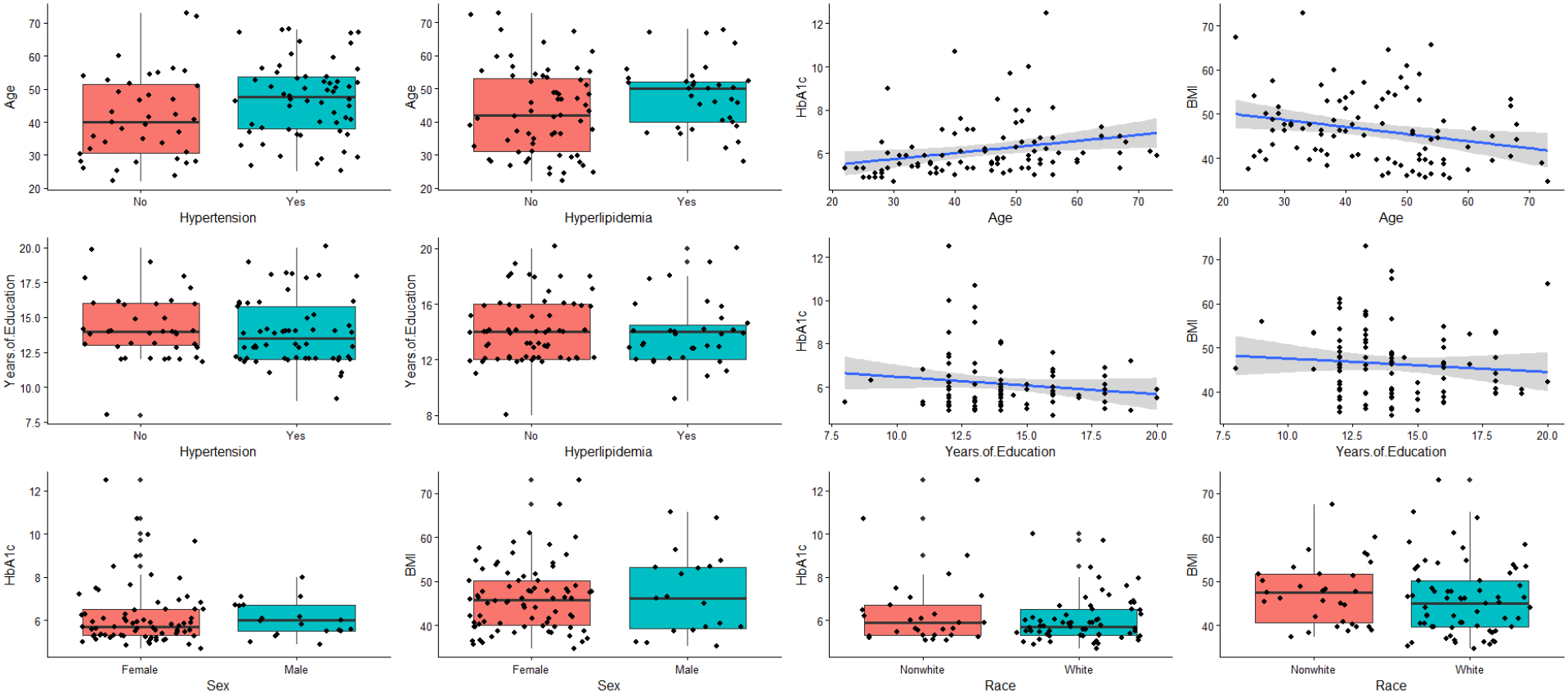
Bivariate Associations of Cardiometabolic Risk Factors and Demographic Variables. Scatterplots and boxplots for hypertension and age, hyperlipidemia and age, HbA1c and age, BMI and age, hypertension and years of education, hyperlipidemia and years of education, HbA1c and years of education, BMI and years of education, HbA1c and sex, BMI and sex, HbA1c and race, and BMI and race.

Due to associations with cardiometabolic risk factors and expected associations with BAG, age and race were added as covariates in multiple linear regression models. Years of education was excluded from linear models due to insignificant associations with cardiometabolic risk factors. Sex was included in linear regression models due to the sex specificity of the ENIGMA brain age prediction algorithm.

### 3.3 Linear Models Assessing Cardiometabolic Risk Factor Associations with BAG

Linear models including HbA1c, BMI, hyperlipidemia, hypertension, age, sex, and race as covariates showed a relationship between HbA1c and BAG for the ENIGMA (BAG_ENIGMA_) and Pyment (BAG_Pyment_) models, such that individuals with higher HbA1c had higher BAG. Statistical characteristics for the Pyment and ENIGMA linear models are included in **Table 4**. BAG_ENIGMA_ was inversely associated with age, consistent with previously reported underestimation of brain age in older individuals.^16^ Accordingly, age was included as a covariate in further analyses. Male sex was associated with increased BAG_Pyment_, but not BAG_ENIGMA_ compared to female participants. No other covariates were associated with BAG for either model. **Supplementary Figures 2 and 3** show added variable plots visualizing fitted partial regression lines for each covariate and BAG after adjusting for the remaining predictors, for BAG_ENIGMA_ and BAG_Pyment,_ respectively.

**Table 4:**
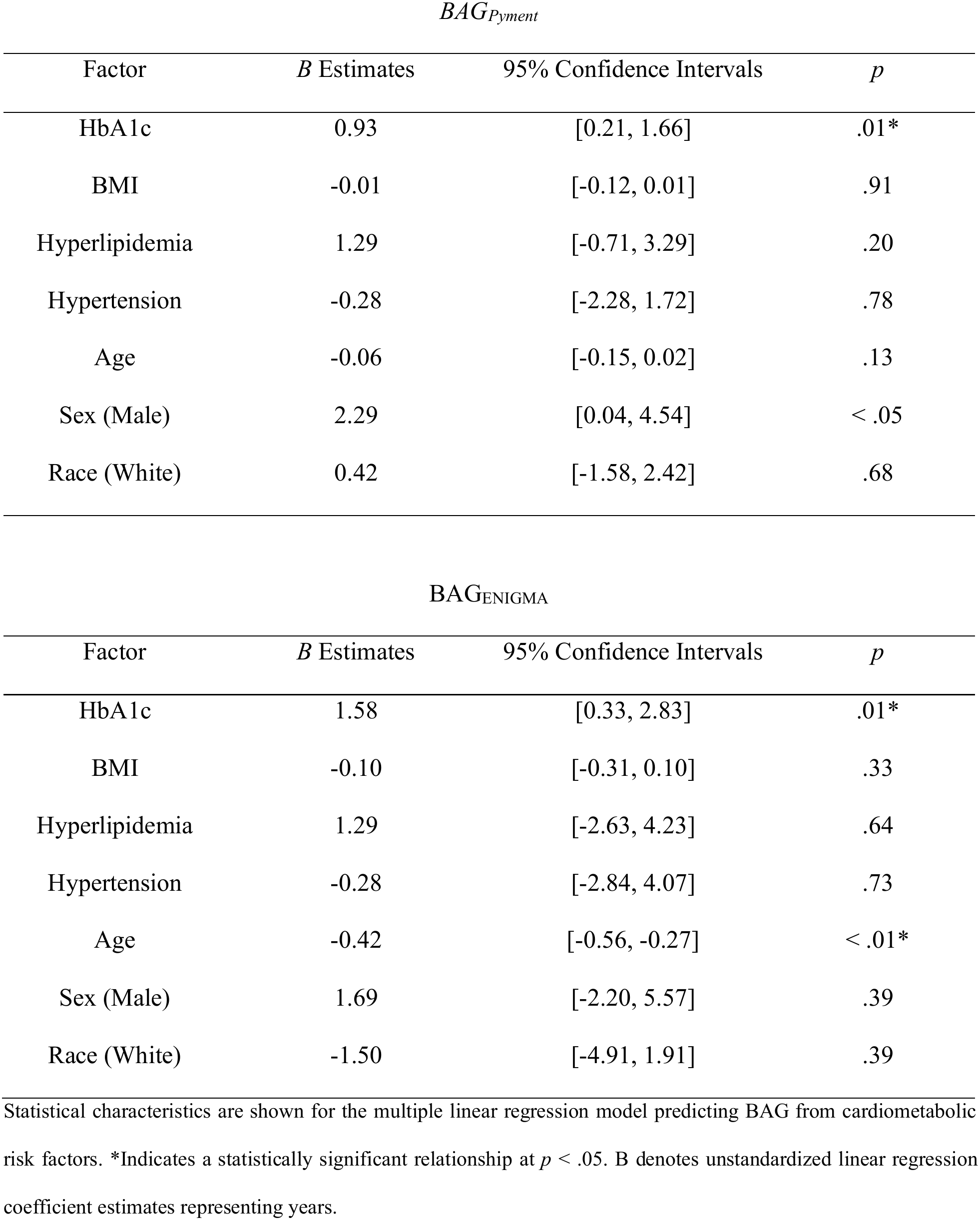
Multiple Linear Regression for BAG_Pyment_ and BAG_ENIGMA_.

| *BAG<sub>Pyment</sub>* |  |  |  |
|---|---|---|---|
| Factor | *B* Estimates | 95% Confidence Intervals | *p* |
| HbA1c | 0.93 | [0.21, 1.66] | .01* |
| BMI | -0.01 | [-0.12, 0.01] | .91 |
| Hyperlipidemia | 1.29 | [-0.71, 3.29] | .20 |
| Hypertension | -0.28 | [-2.28, 1.72] | .78 |
| Age | -0.06 | [-0.15, 0.02] | .13 |
| Sex (Male) | 2.29 | [0.04, 4.54] | < .05 |
| Race (White) | 0.42 | [-1.58, 2.42] | .68 |
| BAG<sub>ENIGMA</sub> |  |  |  |
| Factor | *B* Estimates | 95% Confidence Intervals | *p* |
| HbA1c | 1.58 | [0.33, 2.83] | .01* |
| BMI | -0.10 | [-0.31, 0.10] | .33 |
| Hyperlipidemia | 1.29 | [-2.63, 4.23] | .64 |
| Hypertension | -0.28 | [-2.84, 4.07] | .73 |
| Age | -0.42 | [-0.56, -0.27] | < .01* |
| Sex (Male) | 1.69 | [-2.20, 5.57] | .39 |
| Race (White) | -1.50 | [-4.91, 1.91] | .39 |

Statistical characteristics are shown for the multiple linear regression model predicting BAG from cardiometabolic risk factors. \*Indicates a statistically significant relationship at *p* < .05. B denotes unstandardized linear regression coefficient estimates representing years.

### 3.4 Robust Regression Assessing Cardiometabolic Risk Factor Associations with BAG

MM-Estimation robust linear models were run due to presence of outliers and high leverage points, which are visualized by Cooks Distance and Leverage plots in **Supplementary Figure 1**. In robust linear regression models, HbA1c and age were associated with BAG_ENIGMA_ such that individuals with higher HbA1c and lower age had higher BAG. HbA1c was no longer significantly associated with BAG_Pyment_. No covariates in the model were significantly associated with BAG. Statistical characteristics are shown in **Table 5**. **Figure 2** visualizes marginal effects plots for HbA1c in the robust linear regression models for Pyment and ENIGMA algorithms, respectively.

**Table 5.** MM-Estimation Robust Linear Model for BAG_Pyment_ and BAG_ENIGMA_.

| BAG<sub>Pyment</sub> |  |  |  |
| Factor | *B* Estimates | 95% Confidence Intervals | *p* |
|---|---|---|---|
| HbA1c | 0.71 | [-0.21, 1.63] | .13 |
| BMI | -0.00 | [-0.11, 0.11] | .99 |
| Hyperlipidemia | 1.15 | [1.48, 3.79] | .39 |
| Hypertension | 0.00 | [-1.75, 1.76] | .99 |
| Age | -0.04 | [-0.13, -0.04] | .32 |
| Sex (Male) | 2.09 | [-0.41, 4.60] | .10 |
| Race (White) | 0.33 | [-1.50, 2.16] | .72 |
| BAG<sub>ENIGMA</sub> |  |  |  |
| Factor | *B* Estimates | 95% Confidence Intervals | *p* |
| HbA1c | 1.70 | [0.65, 2.76] | < .01* |
| BMI | -0.08 | [-0.30, 0.14] | .45 |
| Hyperlipidemia | 1.19 | [-1.93, 4.31] | .45 |
| Hypertension | 0.33 | [-3.04, 3.71] | .84 |
| Age | -0.41 | [-0.55, -0.29] | < .01* |
| Sex (Male) | 1.66 | [-2.19, 5.51] | .38 |
| Race (White) | -1.44 | [-4.67, 1.79] | .39 |

\*Indicates a statistically significant relationship at *p* < .05. B denotes unstandardized linear regression coefficient estimates representing years.

**Figure 2.**
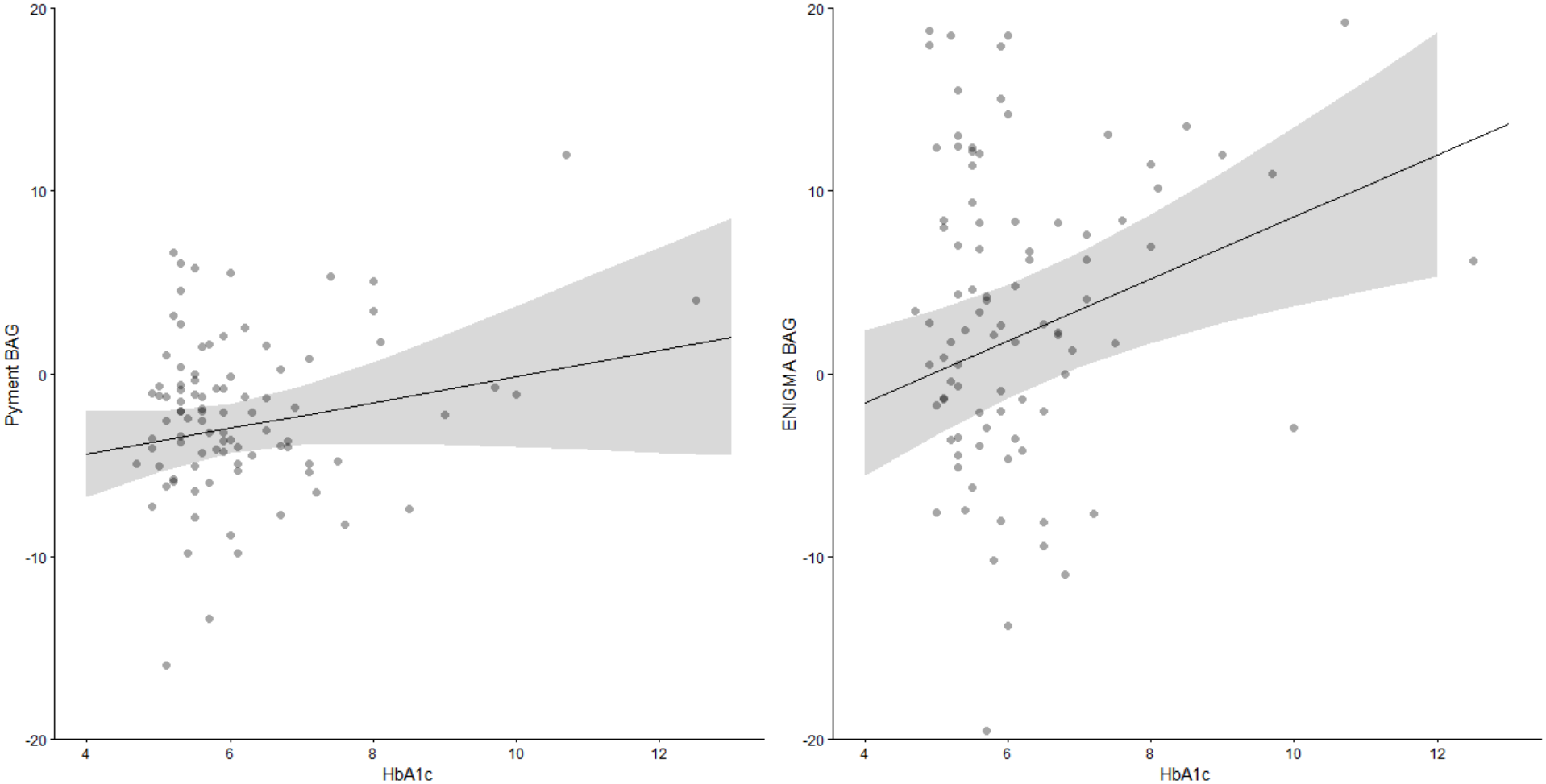
HbA1c and BAG Robust Regression Marginal Association Marginal effects plot showing the model–estimated association between HbA1c and brain age gap (BAG), adjusted for covariates for both Pyment and ENIGMA models on the left and right, respectively. The solid line represents the predicted BAG across the observed range of HbA1c values, and the shaded ribbon reflects the 95% confidence interval. Greater BAG indicates advanced biological brain aging.

### 3.5 Bland Altman Test and Brain Age Correlations with Age

A Bland-Altman analysis between BAG_ENIGMA_ and BAG_Pyment_ showed a statistically significant difference between BAG values for the different algorithms, with BAG_ENIGMA_ values systematically higher than BAG_Pyment_ values (*μ_d_* = 6.10, SD = 8.37, *p* < .001). The limits of agreement ranged from –10.31 years to 22.50 years, indicating relatively wide agreement between models. The lack of agreement between the models is consistent with findings from **Table 1** showing the ENIGMA model overestimating age and the Pyment model underestimating age. **Figure 2** shows the Bland Altman test and **Supplementary Figure 4** shows scatterplots for brain-predicted age and chronological age for ENIGMA (*ρ* = .73, *p* < .001) and Pyment (*ρ* = .92, *p* < .001) algorithms. **Supplementary Figure 5** shows a scatterplot visualizing the relationship between BAG_ENIGMA_ and BAG_Pyment_.

**Figure 3.**
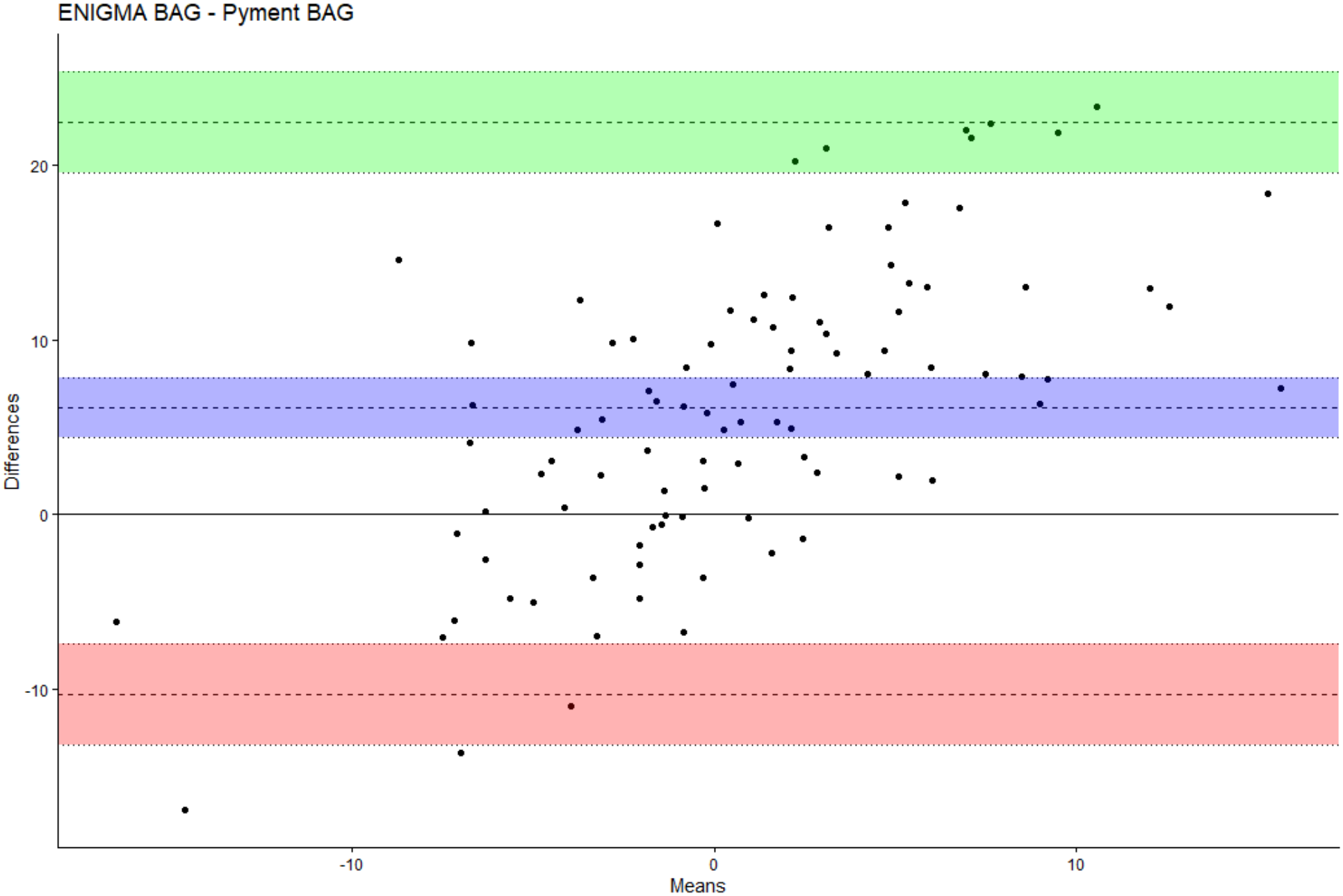
Bland Altman Plot for BAG_ENIGMA_ and BAG_Pyment_ Green, purple, and red areas indicate upper limit of agreement, mean difference, and lower limit of agreement confidence intervals, respectively. As predicted age increases, BAG_ENIGMA_ values are greater in comparison to BAG_Pyment_.

### 3.6 Stratification by Diabetic Status

Post hoc model stratified by diabetic status using both the American Diabetes Association cutoffs for diabetes and prediabetes.^34^ When stratifying by glycemic status using HbA1c, 1 percent increases in HbA1c in normoglycemic individuals were associated with 2.01 and 5.15 year increases on average for BAG_Pyment_ and BAG_ENIGMA,_ respectively, but neither association reached significance (*p* = .42 and *p* = .36). However, in the prediabetes cutoff group, 1 percent increases in HbA1c were associated with an average of 1.21 and 2.15 year increases in BAG_Pyment_ and BAG_ENIGMA_, respectively, with both models showing significant associations (*p* = 0.04 and *p* = .01), and in the diabetes cutoff group, 1 percent increases in HbA1c were associated with an average of 1.12 and 2.49 year increases in BAG_Pyment_ and BAG_ENIGMA_, respectively, with a significant association for BAG_ENIGMA_ (*p* < .05), but not BAG_Pyment_ (*p* = .17). **Table 6** shows the number of individuals, B estimates, p-values, and confidence intervals for each cardiometabolic risk factor for both brain age models in the normoglycemic, prediabetic, and diabetic groups.

**Table 6.** Robust Linear Models Stratified by Diabetic Status.

| Normoglycemic (N = 43) |  |  |  |  |  |  |
|---|---|---|---|---|---|---|
| Algorithm | BAG<sub>Pyment</sub> |  |  | BAG<sub>ENIGMA</sub> |  |  |
| Characteristics | *B* | *p* | 95% CI | *B* | *p* | 95% CI |
| HbA1c | 2.01 | .42 | [-3.02, 7.05] | 5.15 | .36 | [-6.16, 16.47] |
| BMI | 0.01 | .42 | [-0.07, 0.10] | -0.03 | .86 | [-0.37, 0.31] |
| Hyperlipidemia | 4.98 | .001* | [2.09, 7.87] | 2.61 | .40 | [-3.56, 8.78] |
| Hypertension | 0.29 | .80 | [-2.04, 2.62] | -0.49 | .87 | [-6.39, 5.41] |
| Prediabetes/Diabetes (N = 50) |  |  |  |  |  |  |
| Algorithm | BAG<sub>Pyment</sub> |  |  | BAG<sub>ENIGMA</sub> |  |  |
| Characteristics | *B* | *p* | 95% CI | *B* | *p* | 95% CI |
| HbA1c | 1.21 | .04* | [0.15, 2.26] | 2.15 | .01* | [0.51, 3.81] |
| BMI | -0.03 | .80 | [-0.27, 0.21] | -0.19 | .24 | [-0.54, 0.16] |
| Hyperlipidemia | -0.56 | . 71 | [-3.36, 2.24] | 0.33 | .89 | [-4.73, 5.39] |
| Hypertension | 0.56 | .66 | [-1.98, 3.11] | 1.33 | .65 | [-4.59, 7.24] |
| Diabetes (N=26) |  |  |  |  |  |  |
| Algorithm | BAG<sub>Pyment</sub> |  |  | BAG<sub>ENIGMA</sub> |  |  |
| Characteristics | *B* | *p* | 95% CI | *B* | *p* | 95% CI |
| HbA1c | 1.12 | .17 | [-0.53, 2.79] | 2.49 | < .05* | [0.05, 4.94] |
| BMI | 0.03 | .82 | [-0.27, 0.34] | -0.07 | .68 | [-0.41, 0.28] |
| Hyperlipidemia | -0.25 | .95 | [-9.06, 8.56] | 1.30 | .72 | [-7.17, 8.06] |
| Hypertension | -1.44 | .56 | [-6.62, 3.74] | -2.02 | .50 | [-7.93, 4.02] |

\*Indicates a statistically significant relationship at *p* < .05. B denotes unstandardized linear regression coefficient estimates representing years.

## 4. DISCUSSION

In this study, we investigated the relationship between cardiometabolic risk factors, including HbA1c, and a machine-learning approach to estimating biological aging of the brain, in individuals who have severe obesity. We used the difference between estimated brain age and chronological age (BAG) as evidence of advanced brain aging. We assessed whether previously observed associations between brain age gap and cardiometabolic risk factors generalize to a sample with severe obesity.^27–30^ To our knowledge, only one other study has assessed the relationship between brain age gap and cardiometabolic risk factors in populations of individuals with severe obesity.^30^ Zeighami et al.^30^ found associations between brain age gap and higher insulin resistance (HOMA-IR), higher BMI, and higher blood pressure, but they did not test the association using multiple or open-source brain age metrics and only included populations undergoing bariatric surgery.^30^ Our study expands on this previous work evaluating these relationships using two widely used open-source brain age metrics, which rely on distinct structural measurements to estimate brain age. This allows for greater reproducibility of results and the potential for combining results across datasets due to the ability to use the same brain-age algorithms. Unlike prior studies, the sample used in this study also includes individuals with severe obesity from the community who did not undergo bariatric surgery. We believe these results may be more likely to generalize to larger populations with severe obesity including those not undergoing bariatric surgery.

Our study provides evidence to support a relationship between altered glycemic regulation and advanced brain aging, extending upon previously reported relationships between elevated HbA1c and increased brain age gap in populations with severe obesity.^30^ Linear and robust linear models found associations between greater HbA1c and greater BAG_ENIGMA_, while the linear model, but not robust linear model, found associations between greater HbA1c and BAG_Pyment_. When stratifying for glycemic status, both the Pyment and ENIGMA robust linear regression models found associations between HbA1c and BAG in prediabetic individuals. In the ENIGMA model only, an association between HbA1c and BAG was found in diabetic individuals.

The lack of an association between HbA1c and BAG_Pyment_, but association of HbA1c and BAG_ENIGMA_ in robust models could be due to differences in accuracy between the algorithms in clinical populations. The Bland Altman test supported evidence of lack of agreement between separate brain age models, concluding significant systematically higher estimation of brain age for the ENIGMA model than the Pyment model. Previous literature assessing the validity and inter-method reliability of open-source brain age algorithms suggest tighly-fitted algorithms, which had the lowest MAE and highest Pearson’s r among algorithms in healthy testing datasets, may be less indicative of brain aging pathology in clinical populations than less tightly-fitting models, with higher MAE and Pearson’s r in healthy testing populations, with the caveat that there may be more noise in the looser-fitting models.^15^ Therefore, models that are reported as moderate-fitting (MAE ∼6) may be most appropriate when using open-source models to generate brain-predicted age as a biomarker of brain health in individuals with severe obesity, rather than models with the lowest MAE values.

Bashyam et al. found that moderate fitting models found significantly higher differentiation compared to tightly-fitting models in clinical Alzheimer’s and MCI groups and reported their moderate and tightly-fitting models as having mean absolute error (MAE) values of 5.922 and 3.701, respectively.^15^ Our results may support a similar pattern in clinical groups with severe obesity, as the tightly-fitting Pyment model (reported MAE = 3.9, *r* = .975 in healthy testing data) estimated an average age about two years younger than the actual age of participants, while the less tightly-fitting ENIGMA model (reported MAE = 6.50, *r* = .85 for males, MAE = 6.85, *r* = .85 for females in healthy-participant testing data) estimated an average age of about four years older than the true mean age of participants.^17,37^ The ENIGMA estimation indicating greater biological aging in populations with obesity compared to healthy controls is aligned with previous literature reporting pathological structural and cognitive changes that occur in adults with obesity.^26–30^ Therefore, it was likely the more valid model for using brain age gap as a brain health biomarker. The lack of agreement between Pyment and ENIGMA models may also be due to differences in the training samples between algorithms. The Pyment model was largely trained on a subset of UK Biobank MRI data, which included many more individuals overall, and Leonardsen et al. did not report exclusion of participants from training data based on diabetic or metabolic status.^37^

The ENIGMA model was trained primarily on explicit “control” populations (either recruited specifically as healthy controls or drawn from clinical studies that routinely excluded individuals with chronic conditions such as diabetes), resulting in training data that may reflect healthier (and likely biologically “younger”) brains than were included in the Pyment training data^17,37^. This implies that the ages estimated for the ENIGMA model may reflect the biological brain age of idealized “healthy” individuals, while the ages estimated for the Pyment model reflect more “normal” brain structure for a given age. It is notable that while the individuals in the training dataset for the Pyment model did not exclude individuals for cardiometabolic conditions, the UK biobank data from which this dataset was created has been reported to have an overall healthier population than the rest of the UK population, indicative of healthy volunteer effects.^43^ Therefore, while less healthy than the ENIGMA model training dataset, the Pyment model may also not be a true brain health biomarker for normative aging. In the context of populations with obesity, our study supports that less tightly-fitting models and models that use “healthier” training data may be more appropriate when assessing biological brain aging and may be more sensitive to changes in biological brain aging due to pathological brain changes.

This study suggests that the association between blood glucose and advanced biological brain aging occurs principally when blood glucose exceeds clinical thresholds (i.e. prediabetes and diabetes). Post hoc MM–estimation robust regression testing for associations between BAG and categorical HbA1c, adjusting for BMI, hypertension, and hyperlipidemia, age, sex, and race were also run using HbA1c cutoffs of 5.7, 6.5, and 8 for prediabetes, diabetes, and uncontrolled diabetes, respectively. These analyses also supported this conclusion, though only significant associations were found for the uncontrolled diabetes group using the ENIGMA algorithm (B = 7.99, P = .0004). The analysis indicated a higher threshold but larger effect size of the association between HbA1c and brain age gap in individuals with severe obesity. The results of these analyses are shown in **Supplementary Table 1**. The lack of association of HbA1c and BAG for normoglycemic individuals, but associations in prediabetic and diabetic individuals, as well as increased effect sizes for HbA1c-BAG relationships in prediabetes and diabetes compared with the whole sample, may reflect a relationship in which elevated blood glucose may only be associated with biological brain aging in individuals with severe obesity once it reaches higher levels. However, these differences could also reflect increased resilience to accelerated brain aging among individuals with severe obesity without prediabetes or diabetes. Overall, these findings also highlight the importance of monitoring and controlling blood glucose levels in populations with severe obesity and are in line with previous findings suggesting maintaining blood glucose below prediabetic and diabetic levels are associated with better brain health outcomes.^44, 45^

This study provides little evidence to support a relationship between hyperlipidemia and biological brain aging in individuals with severe obesity. There was only one significant association found between hyperlipidemia and BAG, which was a significant association between BAG_Pyment_ and hyperlipidemia in normoglycemic individuals. This is likely reflective of the inclusion criteria for the sample, as the individuals who are normoglycemic, but qualify for bariatric surgery, may qualify due to cardiovascular conditions. They are, therefore, more likely to have more severe cardiovascular conditions. This may drive associations with hyperlipidemia and fail to generalize to other populations with severe obesity.

The study also does not provide evidence to support an association between BMI and BAG or hypertension and BAG in this population with severe obesity, despite significant associations found in a previous study with this population and previous studies investigating relationships in other populations.^26–30^ The lack of association between BMI and BAG in this sample is likely due to selection for high BMI in this sample. There may be a point at which additional increases in BMI correspond to progressively smaller increases in BAG, eventually resulting in no observable association. This pattern would reflect a nonlinear pattern, where the relationship between BMI and BAG is present at lower BMI levels but becomes saturated at higher levels, diminishing the strength of the association.

Some limitations from this study must be considered. Firstly, this study used self-report hyperlipidemia and hypertension data. The self-report data in this study may misclassify some participants, which may contribute to deviations from associations between hypertension, hyperlipidemia, and brain health biomarkers found in other samples with severe obesity. The use of binary variables, as opposed to continuous variables for cholesterol and hypertension, also resulted in lower statistical power and therefore reduced ability for the analyses to find smaller associations with brain age gap if they exist.^46^ Another limitation of this study is its cross-sectional design, which prevents conclusions about causality or the directionality of associations between cardiometabolic risk factors and BAG Longitudinal data would also be necessary to assess how these relationships evolve over time, and to assess whether interventions changing status or level of cardiometabolic risk factors may preserve brain health in individuals over time, as they age. This study also lacks data on the history of cardiometabolic risk factors prior to the visit, such as the duration for which a participant’s HbA1c levels exceeded the diabetes diagnostic threshold. This likely introduces noise into the data or creates systematic bias, leading to difficulty in detecting associations between cardiometabolic risk factors and BAG. Without information on the duration, severity, or clinical measurements of these conditions, the variability in how they manifest across individuals may obscure true relationships.

Future research should incorporate more precise clinical measurements of cardiometabolic risk factors to improve sensitivity and interpretability. Specifically, collecting systolic and diastolic blood pressure values, as well as LDL and HDL cholesterol levels, would allow for a more nuanced understanding of how cardiovascular health relates to brain aging. Future research could also implement cognitive testing to understand the accuracy of the ENIGMA and Pyment brain age algorithms in estimating cognition, in addition to the biological age, in individuals. Although brain age has been shown to correlate highly with cognitive measures in several healthy and clinical populations, and even outperform them in predicting disease outcomes, more evidence for a relationship between brain age and cognitive testing measures in individuals with severe obesity could clarify the relationship between the moderate versus tightly-fitting brain age models and cognition in this population.^30,47^ Additionally, longitudinal studies are needed to track changes in BAG over time and determine whether cardiometabolic interventions can preserve brain health. Investigating the effects of treatments such as bariatric surgery, GLP-1 receptor agonists, behavioral and lifestyle interventions, and medications like statins or antihypertensives may reveal whether improving metabolic health can slow or reverse biological brain aging in individuals with severe obesity and prevent adverse outcomes such as mild cognitive impairment and neurodegenerative disease as they age.

## 5. CONCLUSION

Our work investigating cardiometabolic risk factor associations with brain age gap in individuals with severe obesity shows evidence of an association between elevated HbA1c and advanced brain aging, principally in the context of clinically elevated levels. Interestingly, elevated BMI and self-reported lifetime history of hypertension and hyperlipidemia were not significantly associated with differences in brain age gap. We also identified systematic differences in two open-source algorithms we used to generate brain age gap, along with evidence to support that moderate fitting models and those with training samples that do not include obesity-related comorbidities (e.g. diabetes) may be the most appropriate for use in populations with severe obesity. Further research incorporating longitudinal analyses and characterization of cardiometabolic risk using continuous measured variables (e.g. HDL/LDL, blood pressure), would help to better understand the relationships between cardiometabolic risk and brain age gap in those with severe obesity.

## Data Availability

All data produced in the present study are available upon reasonable request to the authors.

