## Supplementary Materials for "Elevated HbA1c is associated with advanced brain age in severe obesity"

**Supplementary Figure 1.** Cook's Distance and Leverage Plots for Linear Models

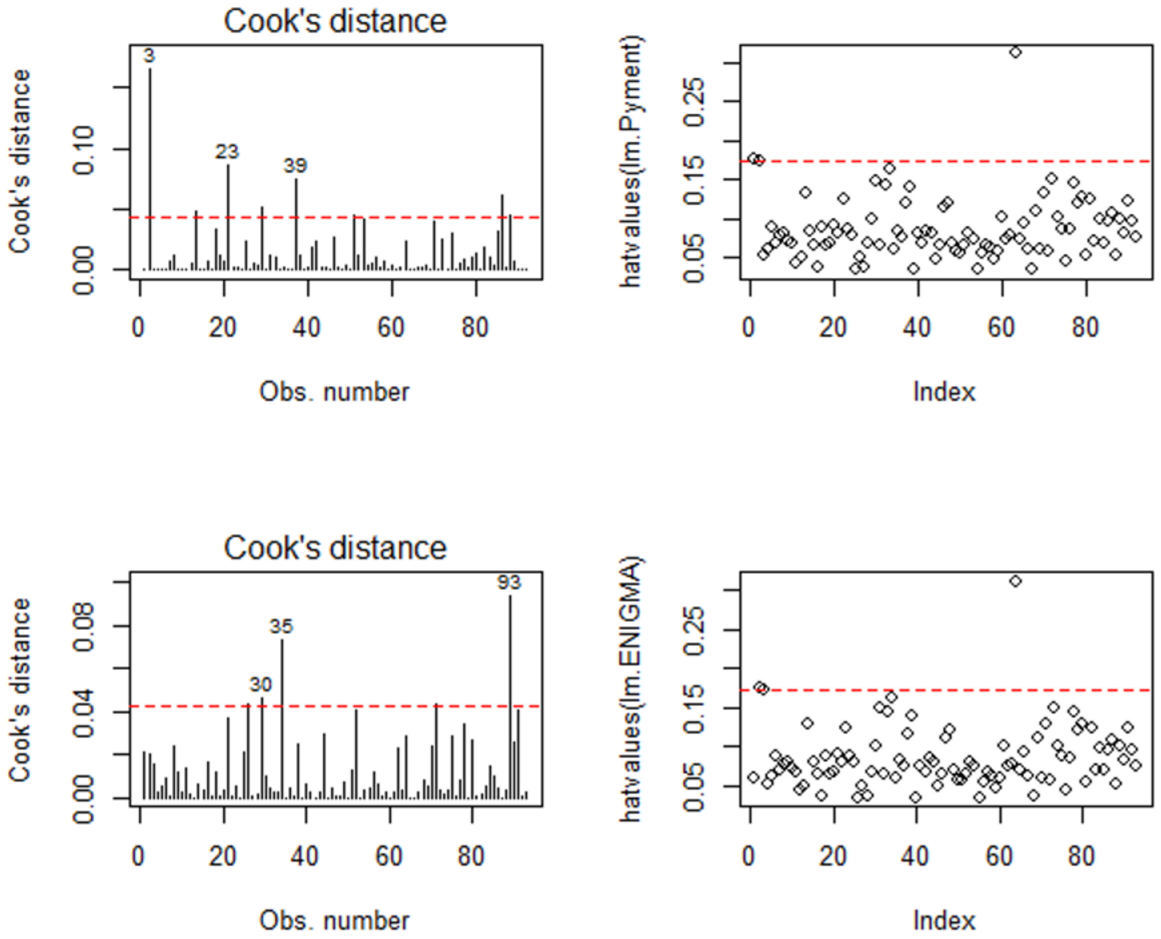


Supplementary Figure 1 shows the Cook's distance (left) and Leverage (right) plots for both the Pyment (top) and ENIGMA (bottom) linear models. Dotted line indicates cutoff values for outliers and high leverage points. Cook's distance plots indicated several outliers (values above 4/n) for both the ENIGMA and Pyment linear models. Leverage plots indicated several high leverage points (values above the 2p/n) for the ENIGMA and Pyment models.

**Supplementary Figure 2.** Added Variable Plots for $BAG_{ENIGMA}$ Linear Model

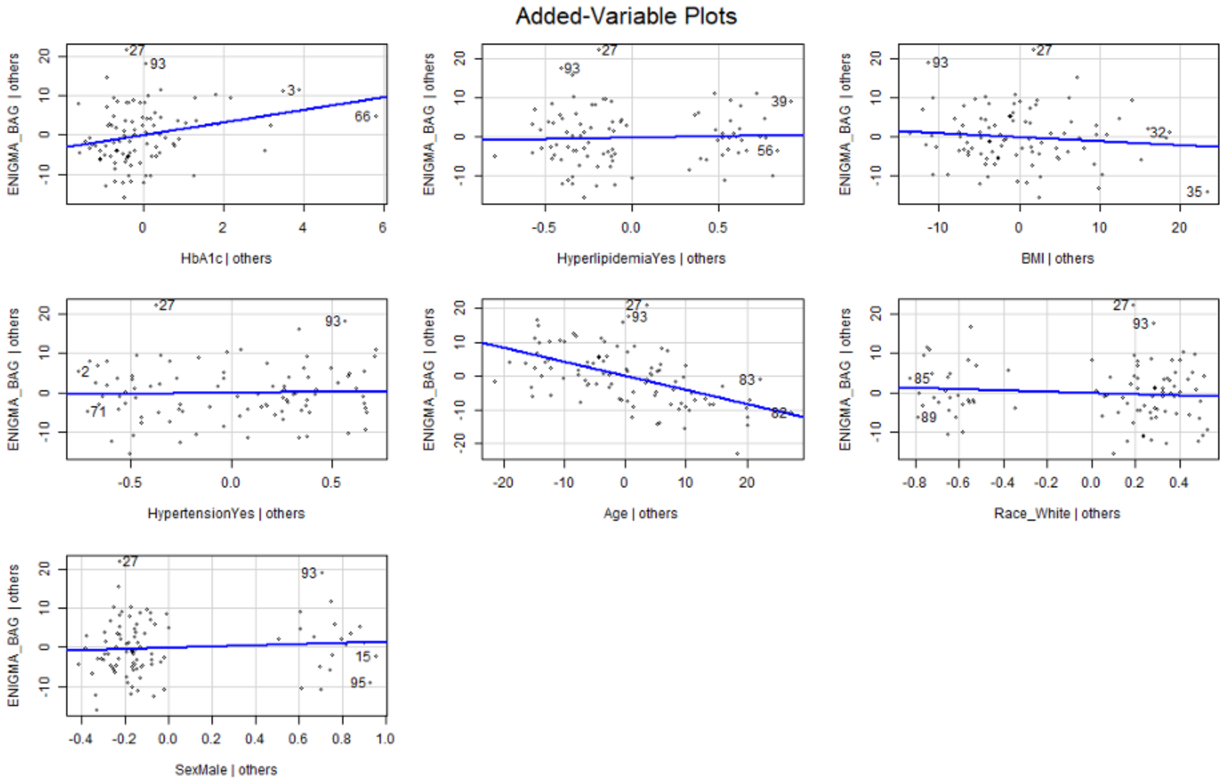


Added variable (partial regression) plots showing fitted partial regression lines between BAG and HbA1c, Hypertension, Hyperlipidemia, Age, BMI, Years of Education, Sex, and Race show the relationship between $BAG_{ENIGMA}$ across individuals.

**Supplementary Figure 3.** Added Variable Plots for $BAG_{Pyment}$ Linear Model

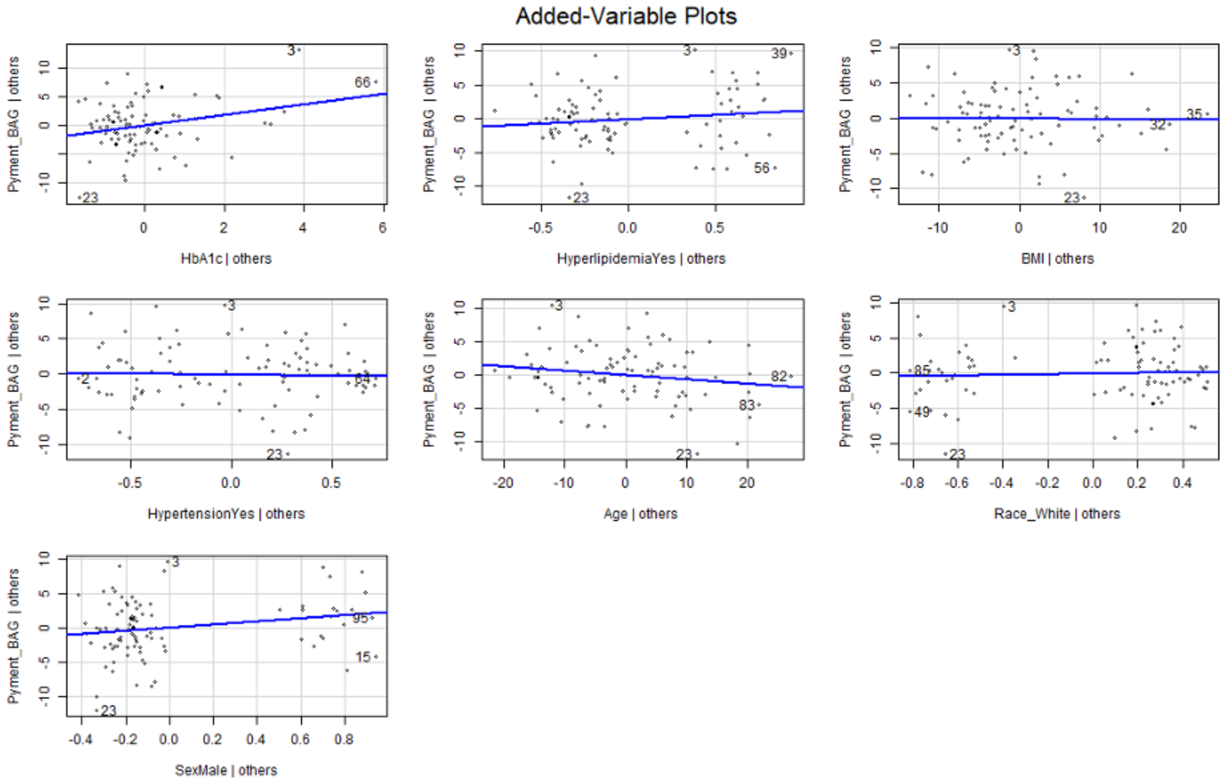


Added variable (partial regression) plots showing fitted partial regression lines between BAG and HbA1c, Hypertension, Hyperlipidemia, Age, BMI, Years of Education, Sex, and Race show the relationship between $BAG_{Pyment}$ across individuals.

**Supplementary Figure 4.** Brain-Predicted Age Correlations with Chronological Age

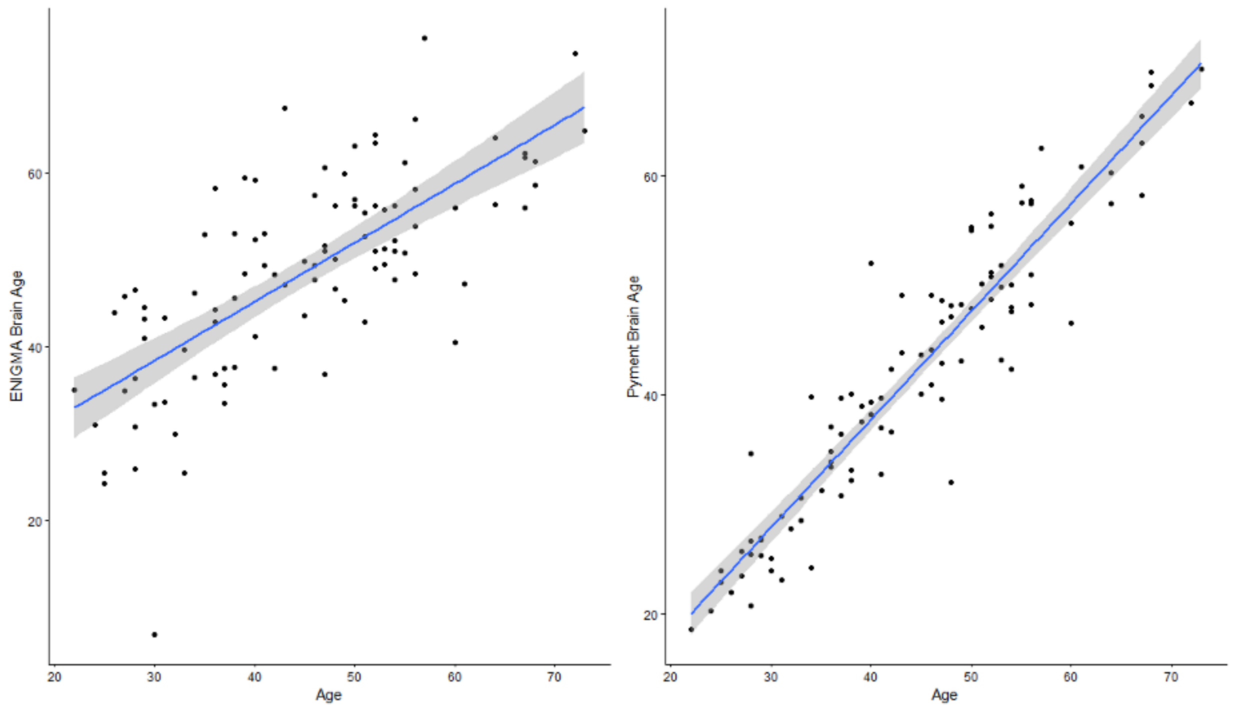


Scatterplots showing line of best fit for associations between brain predicted age and chronological age for ENIGMA ($\rho$ = .73, $p$ < .001) and Pyment ($\rho$ = .92, $p$ < .001) brain age value.

**Supplementary Figure 5.** Pyment Brain Age and ENIGMA Brain Age Scatterplot

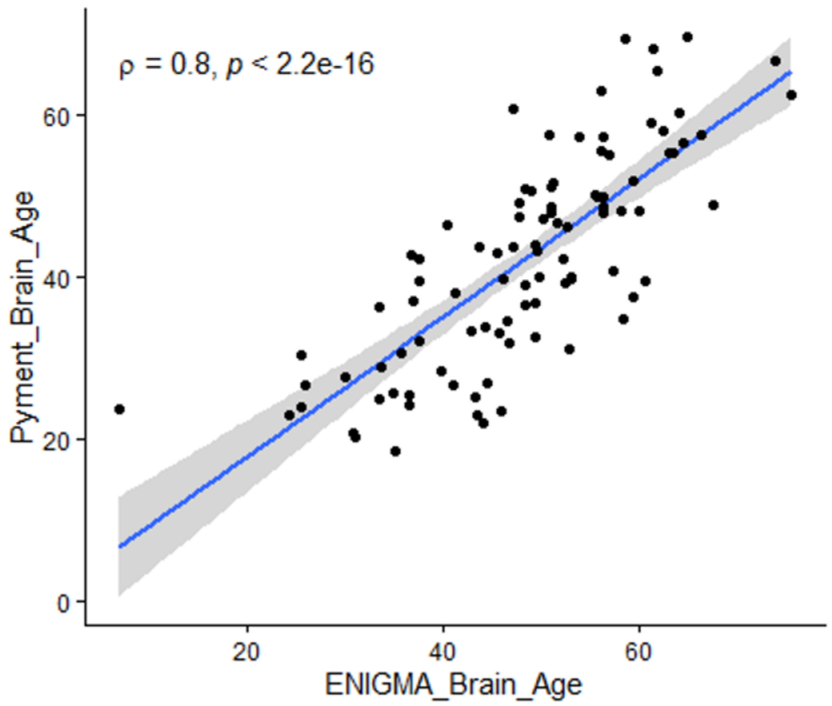

Scatterplot shows the relationship between brain-predicted age across individuals calculated by Pyment and ENIGMA models ($\rho = .80$, $p < .001$).

**Supplementary Table 1.** MM-Estimation Robust Linear Model for $BAG_{Pyment}$ and $BAG_{ENIGMA}$ with Categorical HbA1c

| *BAG<sub>Pyment</sub>* | | | |
|---|---|---|---|
| *Factor* | *B* Estimates | 95% Confidence Intervals | *P* Values |
| *Prediabetes* | -1.04 | [-2.82, 0.74] | .25 |
| *Diabetes* | -1.62 | [-3.93, 0.70] | .17 |
| *Uncontrolled Diabetes* | 3.47 | [-0.09, 7.02] | .056 |
| *BMI* | 0.01 | [-0.09, 0.10] | .89 |
| *Hyperlipidemia* | 1.11 | [-1.06, 3.29] | .31 |
| *Hypertension* | 0.52 | [-1.26, 2.29] | .56 |
| *Age* | -0.02 | [-0.09, 0.06] | .64 |
| *Sex (Male)* | 2.03 | [-0.06, 4.12] | .057 |
| *Race (White)* | 0.30 | [-1.48, 2.04] | .74 |
| *BAG<sub>ENIGMA</sub>* | | | |
| *Factor* | *B* Estimates | 95% Confidence Intervals | *P* Values |
| *Prediabetes* | 0.18 | [ -4.30, 4.67] | .93 |

| | | | |
|---|---|---|---|
| *Diabetes* | 0.09 | [-3.92, 4.10] | .97 |
| *Uncontrolled Diabetes* | 7.99 | [3.65, 12.33] | <.01* |
| *BMI* | -0.04 | [ -0.28, 0.18] | .67 |
| *Hyperlipidemia* | 1.08 | [ -1.85, 4.01] | .47 |
| *Hypertension* | 0.99 | [ -2.67, 4.66] | .59 |
| *Age* | -0.39 | [ -0.52, -0.26] | < .01* |
| *Sex (Male)* | 1.67 | [ -2.74, 6.08] | .33 |
| *Race (White)* | -1.59 | [ -4.80, 1.62] | .45 |

*Indicates a statistically significant relationship. B denotes unstandardized linear regression coefficient estimates representing years. HbA1c was used to categorize diabetes groups into normoglycemic (< 5.7%,) prediabetes (5.7%-6.4%), diabetes (6.5%-8.0%) and uncontrolled diabetes (> 8.0%).
